# Functional MRI Guided Accelerated Intermittent Theta Burst Stimulation for Chronic Post-Concussive Syndrome: A Feasibility and Efficacy Study

**DOI:** 10.1101/2025.10.08.25336734

**Authors:** Evan J. Bartholomeusz, Alina K. Fong, Thomas J. Tervort, Lynn M. Gaufin, Mark D. Allen

**Affiliations:** Cognitive FX, Provo, Utah, USA; Intermountain Healthcare, Salt Lake City, Utah, USA; Utah Neurological Clinic, Provo, Utah, USA

**Keywords:** Post Concussion Syndrome, fMRI Transcranial Magnetic Stimulation, Accelerated Intermittent Theta Burst, Functional Neuroimaging

## Abstract

Post concussive syndrome (PCS) affects 10-15% of people with mTBI (concussion). Some of these symptoms persist for months or years after the original injury. Currently, few effective interventions exist for PCS. Previous studies have explored transcranial magnetic stimulation (TMS) as an interventional measure for PCS. TMS is also demonstrated to be an effective treatment for patients with depression and anxiety, which are often comorbid in patients with chronic concussion-related symptoms. This exploratory study aims to examine the feasibility and efficacy of a rapid, accelerated intermittent theta burst stimulation (iTBS) TMS treatment protocol to alleviate symptoms in patients with chronic PCS.

## INTRODUCTION

Post-Concussive Syndrome is generally characterized by the persistence of a constellation of symptoms following an mTBI, lasting longer than the typical acute recovery period. While diagnostic criteria vary, PCS is often defined operationally by the presence of three or more characteristic symptoms persisting for more than 3 months post-injury. The Diagnostic and Statistical Manual of Mental Disorders, 5th Edition (DSM-5) includes “Neurocognitive Disorder due to Traumatic Brain Injury,” which encompasses persistent cognitive and related symptoms following TBI. It is important to note that PCS is considered a syndrome – a collection of symptoms – rather than a single disease entity, reflecting the heterogeneous nature of its presentation and potentially diverse underlying pathophysiological mechanisms.

Although estimates vary depending on the population studied and diagnostic criteria employed, most head injury cases (70–90%) are classified as mTBI ^1^, and approximately 10-15% of individuals who sustain an mTBI continue to experience debilitating symptoms indicative of PCS for months or even years after their initial injury ^2^. Other studies place the prevalence between 11% and 82% depending on diagnostic criteria. ^3^. Certain factors, such as history of previous concussions, female sex, pre-existing mental health conditions, or litigation status, have sometimes been associated with an increased risk of developing persistent symptoms ^4^, although the interplay of these factors is complex and not fully understood. There is also a significant overlay of psychological symptoms which may confound diagnoses. An overlap exists between post-concussion symptoms and other physical, neurological, and psychiatric conditions, including post-traumatic stress disorder. There are complex interactions between both injury-related and non-injury-related factors, as well as biological, psychological, and social factors, that can lead to post-concussion symptoms ^5^.

Despite the significant burden imposed by Post-Concussive Syndrome (PCS), current standard treatment lacks definitive, universally effective interventions specifically approved for this condition. Management typically relies on a symptom-focused approach aimed at alleviating specific complaints and improving function, rather than targeting a single underlying pathophysiology. Current standard of care may include:

1. Pharmacological Management: Medications are frequently employed to manage individual symptoms. For instance, analgesics (over the counter or prescription) may be used for headaches, antidepressants (like SSRIs or SNRIs) for comorbid mood and anxiety symptoms, specific medications for migraine prophylaxis if indicated, sleep aids for insomnia, and occasionally stimulants for severe fatigue or cognitive slowing. However, pharmacological approaches are often palliative, targeting symptoms rather than root causes. Furthermore, patients with PCS can be sensitive to medication side effects as well as drug-to-drug interactions, and polypharmacy becomes a concern when managing multiple symptoms. There is limited evidence supporting the efficacy of most medications specifically for core PCS symptoms beyond treating comorbidities.
2. Rehabilitative Therapies: This forms a cornerstone of current PCS management. Depending on the patient’s specific symptom profile, referrals may be made for:

- Physical Therapy: To address cervicogenic headaches, neck dysfunction, or balance issues, and to guide a graded return to physical activity, which can be beneficial but requires careful pacing to avoid symptom exacerbation
- Vestibular Therapy: To manage dizziness, vertigo, and gaze instability through specific habituation and adaptation exercises
- Vision Therapy: To address oculomotor problems like convergence insufficiency or accommodative dysfunction, which are common after mTBI
- Cognitive Rehabilitation: Often provided by speech-language pathologists or occupational therapists, focusing on compensatory strategies for memory, attention, and executive function deficits, rather than necessarily restoring baseline function
- Psychotherapy: Cognitive Behavioral Therapy (CBT), Acceptance and Commitment Therapy (ACT), and supportive counseling are often employed to help patients cope with persistent symptoms, manage anxiety and depression, address maladaptive beliefs about recovery, and improve functional engagement.

While the multidisciplinary approach can be helpful for many, significant limitations persist. These include:

- Lack of disease-modifying treatments: Current therapies primarily focus on managing symptoms or providing compensatory strategies. Few therapies, if any, are believed to directly target or reverse the potential underlying neurobiological changes contributing to persistent symptoms (e.g., altered functional connectivity, neuroinflammation, metabolic dysfunction).
- Variable and often incomplete efficacy: Response to current treatments is highly variable among individuals. Many patients experience only partial symptom relief, and a significant proportion remain symptomatic despite engaging in comprehensive rehabilitation programs.
- Symptom overlap and diagnostic complexity: The overlap between PCS symptoms and conditions like primary headache disorders, anxiety, depression, and chronic pain complicates treatment selection and attribution of treatment effects.
- Access, cost, and time commitment: Accessing specialized multidisciplinary care involving various therapists can be challenging due to geographical limitations, insurance coverage issues, and high costs. Furthermore, rehabilitation requires significant time commitment and active participation from the patient.
- Passive vs. active mechanisms: Many current treatments are either passive (medications) or rely heavily on the patient’s ability to learn and implement active compensatory strategies, which can be difficult for those with significant cognitive or aPective symptoms.

These limitations underscore the urgent need for novel therapeutic strategies for PCS, particularly those that might directly modulate the underlying neural circuit dysfunctions hypothesized to contribute to persistent symptoms. Interventions like transcranial magnetic stimulation (TMS), which can non-invasively influence brain activity and plasticity, represent a promising avenue for investigation

A specific patterned form of rTMS, intermittent theta burst stimulation (iTBS), involves delivering short bursts of high-frequency (50 Hz) stimulation repeated at a theta frequency (5 Hz). Preclinical and human studies suggest that iTBS protocols generally induce facilitatory effects on cortical excitability, promoting mechanisms akin to long-term potentiation (LTP) ^6^, a fundamental process involved in learning and memory plasticity. Due to its efficiency (delivering a high number of pulses in a short duration) and its demonstrated potential to induce robust plastic changes, iTBS has gained significant attention, notably receiving FDA approval for the treatment of Major Depressive Disorder ^7^.

The rationale for applying TMS, and specifically iTBS, to PCS stems from converging evidence suggesting that persistent post-concussive symptoms are associated with disruptions in large-scale functional brain networks. Neuroimaging studies in individuals with mTBI and PCS frequently report abnormalities in networks critical for cognition and emotional regulation, including:

- **Default Mode Network (DMN):** Implicated in self-referential thought and introspection, alterations in DMN connectivity (often increased intra-network connectivity or altered connectivity with other networks) are commonly observed post-mTBI and linked to cognitive complaints ^89 10 11^.
- **Cognitive Control Network (CCN) /Central Executive Network (CEN):** Involved in goal-directed attention, working memory, and executive functions, reduced activity or connectivity within the CCN/CEN or between the CCN/CEN and other networks is often associated with cognitive deficits in PCS ^9^
- **Salience Network (SN):** Crucial for detecting relevant internal and external stimuli and switching between the DMN and CCN/CEN, dysfunction in the SN (often involving the dorsal anterior cingulate cortex (dACC) and insula) has been linked to fatigue, affective symptoms, and cognitive difficulties post-concussion ^12 13^.

TMS offers the potential to directly target key nodes within these disrupted networks to normalize pathological activity and connectivity patterns. Excitatory protocols like iTBS could potentially enhance activity in hypoactive regions or networks (like potentially underactive parts of the CCN) or modulate pathological patterns of network interaction.

The anterior prefrontal cortex (aPFC), particularly the middle frontal gyrus of the dorsolateral prefrontal cortex (DLPFC), is a critical hub within the CCN, playing a key role in executive functions, working memory, emotional regulation, and cognitive control – functions frequently impaired in PCS. Furthermore, the DLPFC exhibits significant functional connections with both the DMN (inhibitory) and SN/dACC (excitatory). Targeting this region with excitatory iTBS is hypothesized to enhance cognitive control processes and potentially rebalance aberrant network dynamics. Typical accelerated depression treatment with iTBS involves an analysis of resting-state fMRI in which regions of lDLPFC (typically in rostral middle frontal gyrus [rMFG]) are found to strongly anti-correlated with subgenual anterior cingulate cortex (sgACC). The sgACC is hypothesized to mediate depression symptoms and thus targeted via inhibitory connections from rostral middle frontal gyrus ^7^. However, recent studies evaluating connectivity changes after accelerated depression treatment have not found increased inhibitory connections from rMFG to sgACC, but rather find increased inhibitory strength from rMFG to the entire medial DMN ^14^. Recognizing the inter-individual variability in functional brain organization, especially after injury, this study employed resting-state functional connectivity MRI (fcMRI) to personalize the target location within the aPFC for each participant. Specifically, the target was selected based on its functional relationship with key networks implicated in PCS: seeking a site anti-correlated with the DMN (potentially enhancing healthy modulatory functioning between CNN-DMN) while being positively correlated with the dACC and other CCN nodes (potentially enhancing cognitive control network function).

Traditional rTMS protocols typically involve one session per day over several weeks. Accelerated protocols, administering multiple sessions per day over consecutive days, have shown promise in rapidly reducing symptoms in depression ^7^, potentially by inducing more robust and faster cumulative effects on neuroplasticity. Applying an accelerated iTBS protocol in PCS offers potential advantages in terms of treatment duration and patient convenience, making it an attractive approach for investigation in this population, though its efficacy and tolerability specifically for PCS require systematic evaluation.

Therefore, based on the potential for iTBS to modulate cortical plasticity within key brain networks often disrupted in PCS, combined with personalized fcMRI-guided targeting of the aPFC and the potential efficiencies of an accelerated protocol. This exploratory study aims to evaluate the feasibility and preliminary efficacy of an accelerated intermittent theta burst stimulation (iTBS) protocol in individuals with post-concussion syndrome (PCS). Given the potential of iTBS to modulate cortical plasticity within key brain networks disrupted in PCS—particularly when combined with individualized, functional connectivity MRI (fcMRI)-guided targeting of the anterior prefrontal cortex (aPFC)—this study seeks to determine whether this personalized, accelerated neuromodulation approach can meaningfully reduce symptom burden, enhance cognitive function, and improve overall quality of life in this population.

## METHODS

### Study Design

This exploratory pilot study utilized a single-arm, open-label design. The primary purpose was to assess the feasibility, tolerability, and preliminary efficacy of an accelerated intermittent theta burst stimulation (iTBS) protocol delivered to the anterior prefrontal cortex in participants with chronic Post-Concussive Syndrome (PCS). Outcome measures were assessed at baseline, immediately following the 3-day treatment course, and during a short-term follow-up period approximately 1 to 3 weeks post-treatment. BRANY Institutional Review Board granted this study an exempt status as a secondary review of existing data (Study No. 25-12-393-2253). Given the exploratory nature and lack of a control condition, the findings are intended to inform the design of future, more definitive randomized controlled trials.

### Participants

Participants were primarily recruited from referrals to Neural Effects (an insurance-based acute concussion care center tied to Cognitive FX) who presented with chronic concussion symptoms. Two participants with acute (<3 months since injury) concussion symptoms were also included.

Participants were screened and were excluded for psychological comorbidities (MDD, GAD, bipolar depression, PTSD, schizophrenia) and any physical or medical contraindications (e.g. metal implants in skull, pacemaker, history of seizures or epilepsy) prior to enrollment. Screened participants were also excluded if they had received prior treatment for PCS at Cognitive FX. 17 total participants were initially screened for participation in this study. Of those, 12 were eligible, and 11 chose to enroll and complete treatment. Three initial participants were treated and reported good outcomes. These preliminary participants were administered pre- and post-task-related fMRI scans, without BDI, BAI, or PCSS surveys. The remaining eight participants were treated and administered pre- and post-task-related fMRI scans, BDI, BAI, and PCSS survey measures.

Eleven participants, four male and seven female, with history of concussion/mTBI were recruited for this study. Patients ranged from 16-72 years of age. Participants were screened to ensure compatibility with treatment requirements. Participants had no history of seizures, no metallic implants in the head or neck area, and no major psychological comorbidities. ANT Neuro Visor2 neuronavigation software was used to guide TMS coil placement and treatment administration according to the location determined by Resting –State fMRI data. iTBS was administered to the target site for each patient across 15 sessions, 1200 pulses per session, 30-minute interval between sessions, 5 sessions per day across 3 consecutive days. Target treatment intensity was 120% resting motor threshold. For eight of the participants, emotional symptoms were assessed using a self-reported Beck Depression Inventory (BDI) and Beck Anxiety inventory (BAI), measured prior to, immediately after, and 1-week post-treatment. Cognitive and functional concussion-related symptoms were assessed with a post-concussion symptom scale (PCSS) before and after each day of treatment, as well as 1-week post-treatment. All patients received fNCI scores measuring brain activity and inter-and intra-regional connectivity while performing three different cognitive tasks. Pre-treatment and 1-2 week post-treatment fNCI scores were compared for all participants.

**Table 1:** Characteristics of participants in exploratory pilot study (symptom report measures, n=8)

| Variable | Mean | SD |
| --- | --- | --- |
| Age | 40.38 | 18.98 |
| Time since Injury (months) | 22.93 | 22.23 |
| Baseline PCSS Score | 54.25 | 31.04 |
| Baseline BDI Score | 21.5 | 10.94 |
| Baseline BAI Score | 22.75 | 15.84 |
| rMT | 52.38 | 3.60 |
| Treatment Intensity (%rMT) | 112.48 | 11.07 |
|  | N | % |
| Male | 4 | 50 |

**Table 2:**
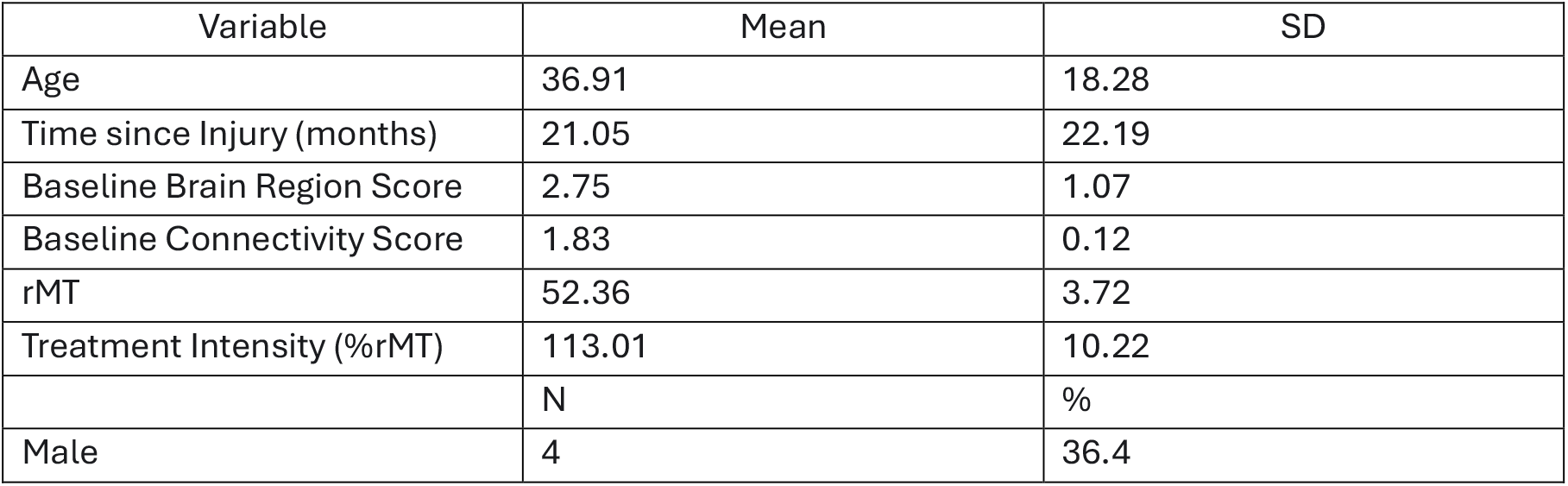
Characteristics of participants in exploratory pilot study (fNCI measures, n=11)

### TMS Procedure

Prior to treatment, patients received a resting state functional connectivity MRI (fcMRI) and a task-related fMRI scan, or functional neurocognitive imaging (fNCI) to examine regional activation and inter-region connectivity, as compared to a database of healthy controls. The purpose of the fcMRI scan was to determine a target location in the anterior prefrontal cortex, anti-correlated with anterior default mode network (DMN) and positively correlated with dorsal anterior cingulate cortex (dACC) and other cognitive control network (CCN) nodes. The purpose of the task-related scan was to provide measures of pre- and post-treatment neurocognitive functioning.

Prior to the first treatment session, each participant’s rMT (resting motor threshold) was determined using the Magstim stimulator equipped with a D70 MT Remote coil (Part No. 4949-00). rMT was defined as the minimum stimulator intensity required to elicit a visible motor response (e.g., twitch in the contralateral abductor pollicis brevis muscle) in at least 5 out of 10 consecutive trials when stimulating the optimal scalp location over the primary motor cortex (M1). This RMT value served as the basis for determining the treatment stimulation intensity.

Intermittent Theta Burst Stimulation (iTBS) was administered using the Magstim Horizon 3.0 (Part No. 5795-00-50) stimulator, equipped with a Horizon 3.0 E-z Cool Coil (Part No. 5620-00-50). The target stimulation intensity was set at 120% of the individual’s determined rMT. Each iTBS session consisted of 1200 pulses delivered in 40 cycles of 10 bursts of 3 pulses at 50 Hz. Each cycle consisted of 2-second trains of bursts and 8 seconds of rest (5Hz).

### Total Pulse and Time Sequence

18,000 total iTBS pulses were administered over 3 consecutive days, with the following exceptions: one participant received 18,000 total iTBS pulses administered on 3 non-consecutive days within a 6-day period, one participant received 15,600 total iTBS pulses administered over 3 consecutive days, and one participant received 15,600 total iTBS pulses administered over 3 non-consecutive days within a 5-day period. Each day of treatment consisted of five sessions of 1200 pulses each, according to the protocol described above. Inter-session intervals each day were 25-35 minutes. All patients received five sessions each day, apart from the two patients who received 15,600 total iTBS pulses. Total treatment time during each session was 6 minutes 32 seconds, for a combined treatment time of 98 minutes.

### Symptom Report Measures

Behavioral measures were assessed using post-concussion symptom scale (PCSS), Beck Depression Inventory (BDI), and Beck Anxiety Inventory (BAI). PCSS is a 21-question Likert scale questionnaire regarding physical, cognitive, sleep, and emotional symptoms of PCS. BDI is a 21-question multiple-choice self-report inventory used for measuring severity of depression. BAI is a 21-question Likert scale questionnaire used for measuring severity of anxiety symptoms.

### Neuroimaging Procedure

#### Task-Related fMRI

Task fMRI data were acquired using a modified version of the Functional Neurocognitive Imaging (fNCI) protocol as described in Epps and Allen^15^. The fNCI assessment protocol combines the validity of conventional neuropsychological testing standards with the reliability and objectivity of informational data output provided by fMRI. The test battery employed in the fNCI was developed and refined through iterative pilot testing to ensure concurrent validity, reliability, objectivity, and suitability for the MRI scanning environment. The current fNCI consists of three tests, a matrix reasoning/visual search test, a picture memory encoding and retrieval test, and a category fluency test. Each test has a duration of 4.5 minutes. Test stimuli were displayed on an MRI compatible CTR screen and test responses were collected using a fiber optic button pad. The tasks were developed with the intent to activate a wide range of functional brain regions and engage a wide range of functional networks, in a manner that is sufficient to detect differences between individuals with no history of significant neurological or psychiatric dysfunction, and those with a history of mild traumatic brain injury. A sample of 270 healthy control volunteers were scanned on the fNCI protocols to provide a normative context to evaluate brain activation levels and functional network connectivity in brain-injured patients. The fNCI is routinely used at Cognitive Fx as a pre-treatment assessment for therapeutic targeting, and as a post-treatment measure of therapeutic efficacy.

#### Resting-State fcMRI

All patients underwent 30 minutes of resting-state fMRI BOLD scanning for the purpose of TMS target location optimization. During the scan, a white fixation cross on a black background was displayed on an MRI compatible CTR screen. Patients were instructed to keep the fixation cross in view during the scan and to let their “minds wander” and to “avoid repetitive thoughts.”

#### Image Acquisition

To provide precise anatomical localization for both the task-related and resting-state analyses, a high resolution T1-weighted MPRAGE image was acquired with the parameters TR = 1645 ms, TE = 3.8 ms, TI = 600 ms, flip angle = 12, 176 slices, FOV = 256 mm. For the task-related and resting-state fMRI scans, 33 axial EPIBOLD images with the critical parameters TR = 2000 ms; TE = 40 ms; Flip Angle = 90 were acquired in inferior to superior interleaved order with a slice thickness of 3 mm, 1 mm interslice gap, with a 64 x 64 matrix for a 3.75 x 3.75 mm in-plane resolution.

#### Image Processing and Analysis

##### Task-Related fMRI

Functional images were preprocessed with a custom pipeline including motion correction and EPI distortion unwarping, ICA image correction, and acquisition time realignment, using sinc interpolation. No head movement exceeded 1 mm translation or 1° rotation displacement. No spatial smoothing was applied.

The objective of the task-related analysis was to assess two critical measures, functional region activation values, and a functional connectivity matrix. These measures were first obtained in the healthy control subjects and organized into a normative brain atlas.

For functional region analysis, the T1-wieghted structural image of each control subject and each patient was warped to the mean functional image after motion correction and unwarping using a custom non-linear coregistration tool. After image alignment, a custom segmentation and functional region parcellation tool (AutoParc) was applied to the T1 images^16^. This cortical labeling protocol uses the “DKT” atlas included in the Mindboggle-101 dataset^17^ providing 31 labels per hemisphere.

The procedure for functional region statistical analysis is described more fully by Lorzel and Allen^16^. A Hidden Markov Model (HMM) process was applied first at the individual voxel level. Further analysis was applied to assess resonant behavior of voxels within a region and passed into a second level HMM to provide whole region activation states. Hemodynamic BOLD activation time series were then assessed with respect to region model states, yielding a t-value for each parcellated region. Individual patient t-values are expressed as z-scores in the context of the t-value normative distribution for each functional region from the control subjects. Anderson–Darling sample-size adjusted tests for normality determined the distributions of each of the 62 regions to be sufficiently normal, with estimates ranging from moderate (A2* = 0.59, p = 0.11) to high (A2* = 0.18, p = 0.91).

Whole-region time-series were assessed for functional connectivity with a non-directed model-free mutual information analysis^16^. Resulting t-values for each inter-region connection from the control subjects were organized into a normative structure, like the procedure for functional regions. Functional connectivity values for patients are reported as z-scores.

#### Resting-State fcMRI

Resting-State fMRI data were processed and analyzed to generate personalized targets in left dorsolateral prefrontal cortex for TMS stimulation. Preprocessing and statistical analysis were performed primarily using tools provided in the FSL software suite (www.fmrib.ox.ac.uk). Preprocessing comprised slice timing correction for interleaved acquisition, motion correction (6-parameter affine transformation) and realignment, spatial smoothing (Gaussian kernel FWHM 5 mm), and temporal high-pass filtering (Gaussian-weighted least-squares straight line fitting, sigma = 100.0 s).

All analyses were done in the patient’s own brain space. T1-weighted structural scans were parcellated using the AutoParc tool and coregistered to the functional scan data space. Non-linear transformation parameters from this warping process were saved to inverse-warp post-analysis activation maps back to structural space. Global signal regression was estimated using the mean functional (EPI) image as a spatial mask. Global signal and head motion parameters were regressed from the BOLD data during calculation of statistical correlational mapping.

For each voxel in functional brain space, correlational values were calculated with a constrained DMN (default mode network) times-series. A standardized DMN mask ^18 19^ (available from https://github.com/yangzhi-psy/PMN-and-DMN-ICA-template) was inverse warped to subject space. The DMN template was generated by averaging the results from 3 group-level ICA algorithms (temporal concatenate ICA, gRAICAR, and independent vector analysis). The DMN mask was constrained by individual anatomical boundaries produced by the AutoParc parcellation to include only medial-prefrontal and orbital frontal portions of the DMN. After correlational analysis, the resulting activation map was inverse warped to patient T1 space.

From the resulting activation map, a single TMS target point (6 mm sphere) was identified by thresholding activation to isolate the peak with the highest t-value. A single unambiguous peak was present in all patients. Target peaks for all patients fell within rostral middle frontal gyrus near the border with inferior frontal gyrus approximately superior to the anterior extent of pars triangularis.

## RESULTS

### FEASIBILITY AND TOLERABILITIY

Commonly reported side effects mirrored results from previous iTBS TMS studies ^7^. Side effects included scalp sensitivity during and immediately after treatment, facial muscle fatigue, headache persisting through inter-treatment intervals, and temporarily worsened self-reported PCSS scores immediately following each treatment session. However, all patients had recovered from side effects prior to beginning the following day’s treatment session.

Due to heterogenous facial nerve distribution across participants, treatment tolerability varied between patients. Treatment was administered at the highest %rMT reported to be tolerable. Accordingly, average treatment intensity across all sessions varied between 98% and 120% rMT, with a mean session treatment intensity of 112.48% rMT. No significant outcome difference was observed in patients who received lower intensity treatment.

### Symptom Report Outcomes

#### PCSS

Mean PCSS scores at three measurement points were as follows, Baseline: 54.25 (SD = 31.04); Post-Treatment: 22.00 (SD = 13.50); 1-4 Weeks Post Treatment: 19.25 (SD = 15.10), see **FIGURE 1**. A one-way ANOVA revealed a significant change in PCSS score over time [F(2,21) = 6.61; p =.006]. Post-hoc independent paired t-test analyses revealed a significant difference between Baseline and Post-Treatment (t = 3.19; p =.008), a significant difference between Baseline and 1-4 Weeks Post Treatment (t = 2.81; p =.013), but no significant difference in the observed trend toward improvement between Post-Treatment and 1-4 Weeks Post Treatment (t <1).

**Figure 1.**
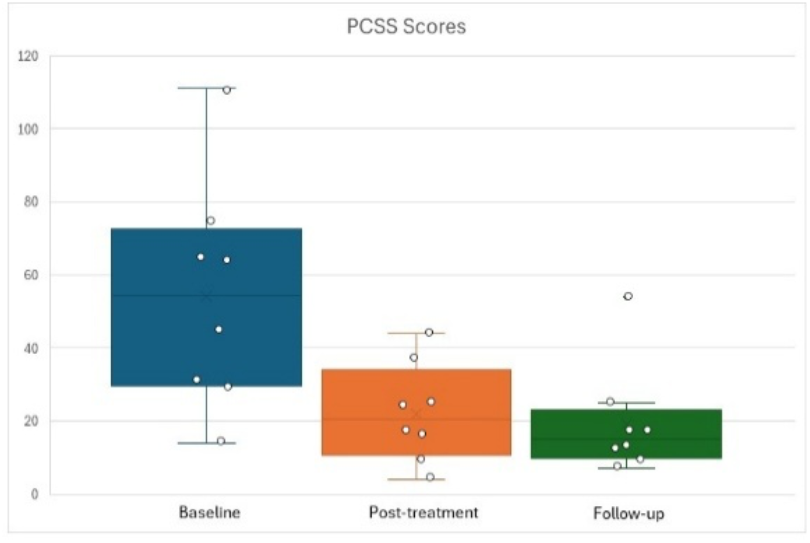
Average PCSS Scores showing significant improvement between Baseline and Post-Treatment (t = 3.19; p =.008); and significant improvement between Baseline and 1-4 Weeks Post-Treatment (t = 2.81; p =.013).

### BDI

Mean BDI scores at three measurement points were as follows, Baseline: 21.5 (SD = 10.94); Post-Treatment: 12.87 (SD = 10.38); 1-4 Weeks Post Treatment: 13.12 (SD = 11.35), see **FIGURE 2**. A one-way ANOVA revealed no overall significant difference in BDI score with respect to time [F(2,21) = 1.62; p =.22]. However, post-hoc independent paired t-test analyses revealed a significant difference between Baseline and Post-Treatment (t = 2.25; p =.03), a significant difference between Baseline and 1-4 Weeks Post Treatment (t = 2.74; p =.014). There was no significant difference between Post-Treatment and 1-4 Weeks Post Treatment (t <1).

**Figure 2.**
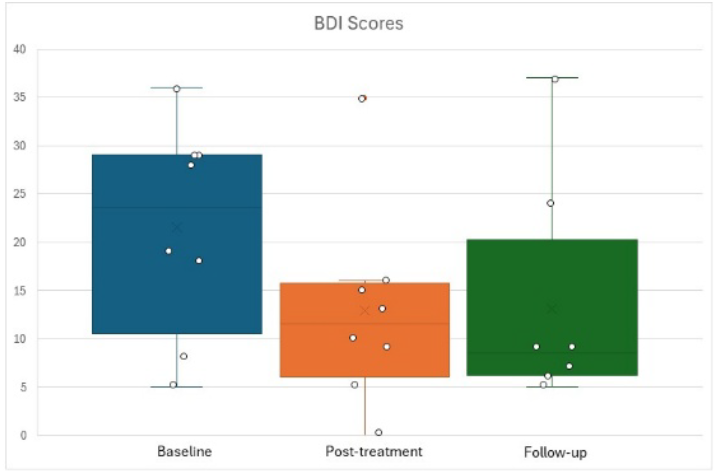
Average BDI Scores showing significant improvement between Baseline and Post-Treatment (t = 2.25; p =.03); and significant improvement between Baseline and 1-4 Weeks Post-Treatment (t = 2.74; p =.014).

### BAI

Mean BAI scores at three measurement points were as follows, Baseline: 22.75 (SD = 15.83); Post-Treatment: 10.62 (SD = 9.37); 1-4 Weeks Post Treatment: 8.62 (SD = 5.78), see **FIGURE 3**. A one-way ANOVA revealed a significant change in BAI score over time [F(2,21) = 3.77; p =.04]. Post-hoc independent paired t-test analyses revealed a significant difference between Baseline and Post-Treatment (t = 1.89; p =.05), a significant difference between Baseline and 1-4 Weeks Post Treatment (t = 3.08; p =.009), but no significant difference in the observed trend toward improvement between Post-Treatment and 1-4 Weeks Post Treatment (t <1).

**Figure 3.**
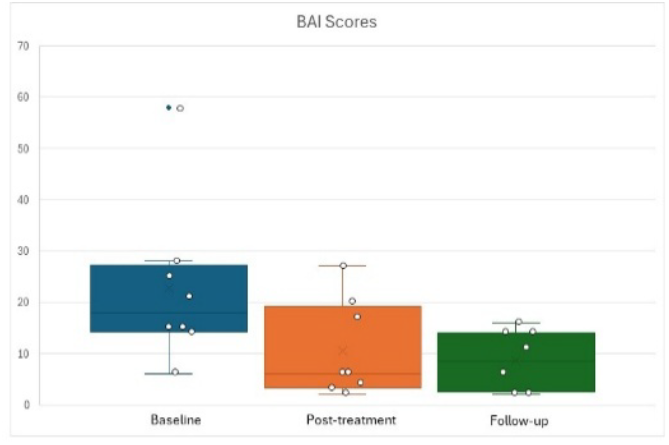
Average BAI Scores showing significant improvement between Baseline and Post-Treatment (t = 1.89; p =.05); and significant improvement between Baseline and 1-4 Weeks Post-Treatment (t = 3.08; p =.009).

**Figure 4.**
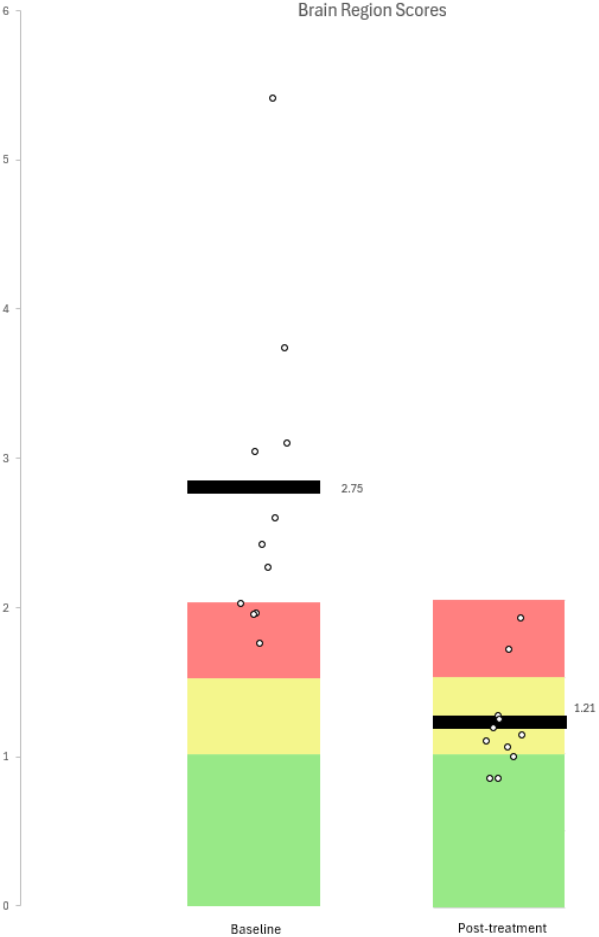
fNCI Brain Region Scores showing significant improvement change from baseline to post-treatment (t = 4.72; p =.0004). Values represent overall deviation from the normative standard, where larger values indicate greater functional impairment. Individual (dots) and average (bar) severity scores are displayed in the context of index normative ranges, where green indicates absence of impaired functioning

**Figure 5.**
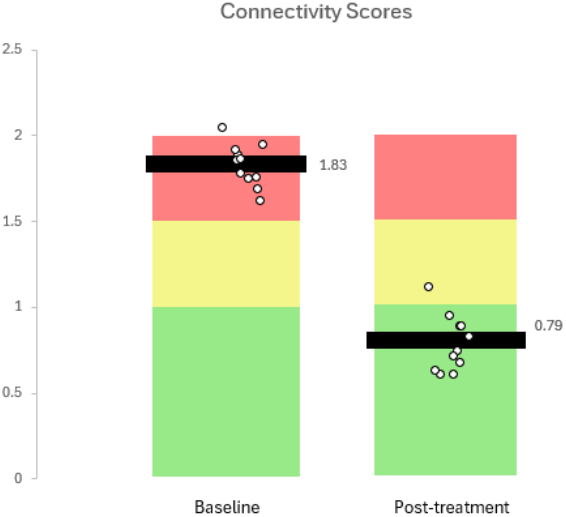
fNCI Functional Connectivity Scores showing significant improvement change from baseline to post-treatment (t = 7.11; p <.0001). Values represent overall deviation from the normative standard, where larger values indicate greater functional impairment. Individual (dots) and average (bar) severity scores are displayed in the context of index normative ranges, where green indicates absence of impaired functioning.

### Neurocognitive Imaging (Task-Related fMRI) Outcomes

fNCI scores showed significant improvements across all participants. Brain Region scores showed an average decrease of 51.6% and Functional Connectivity scores showed an average decrease of 56.9%. For Brain Region scores, where larger values represent greater deviation from the normative standard, the Baseline mean was 2.75 (SD = 1.07), and the Post-Treatment mean was 1.21 (SD = 0.34). For Functional Connectivity scores, the Baseline mean was 1.83 (SD = 0.12), and the Post-Treatment mean was 0.79 (SD = 0.16). Paired t-test analyses revealed a significant difference between Baseline and Post-Treatment for Brain Region scores (t = 4.72; p =.0004), and a significant difference between Baseline and Post-Treatment for Functional Connectivity scores (t = 7.11; p <.0001).

### Summary of Outcomes

Of participants included in the surveyed sample, 6 provided follow-up survey scores at 1-week post-treatment, while 2 provided follow-up survey scores at 4 weeks post-treatment. Participants exhibited a significant (p<0.01) reduction in PCS symptoms immediately following treatment (63% mean symptom reduction, SD=31%), an effect which remained (p<0.02) 1-4 weeks post-treatment (62% mean symptom reduction, SD=42%. Seven participants reported meaningful (>50%) reduction of symptoms immediately following treatment. During the 1-4 week follow-up period, four participants reported a further (>40%) reduction in symptoms, while remaining participants reported little change or moderate increase of symptoms. Patients exhibited a significant (p<0.01) decrease in anxiety symptoms 1-4 weeks post-treatment (60% mean symptom reduction, SD=21%) (14.13 mean BAI score decrease, SD=12.2), but not immediately after treatment. Conversely, patients exhibited a significant (p<0.03) decrease in depression symptoms immediately after treatment (34% mean symptom reduction, SD=40%) (8.63 mean BDI score decrease, SD=10.15) and 1-4 weeks post-treatment (8.38 mean BDI score decrease, SD=8.09).

## DISCUSSION

This pilot study demonstrates that a brief intervention of iTBS TMS, administered over 3 days, is a feasible intervention to alleviate symptoms in patients with chronic PCS and warrants further research. Patients demonstrated significant improvement in task-related brain region activation levels and inter-region functional connectivity compared to healthy controls. Patients also demonstrated significantly reduced self-reported measures of PCS severity, anxiety, and depression symptoms. Improvements across all measures were evident immediately following TMS intervention and persisted over 1-4 weeks after concluding treatment.

Previous findings have shown that chronic PCS symptoms correlate with abnormal functional connectivity in resting state fMRI data ^20^, an observation mirrored in the present pilot study. Data from fNCI assessments suggest that accelerated iTBS TMS rapidly induces metabolic and connective changes within participants’ brains, an effect that persists beyond conclusion of treatment. Participants received fNCI scans ranging from immediately (∼1 hour) post-intervention to approximately 4 weeks post-intervention. All participants demonstrated significant (≥40%) reductions in functional connectivity scores, regardless of time between treatment and follow-up scan.

Application of accelerated iTBS TMS resulted in rapid symptom reduction in all but one patient. Most participants reported substantial symptom abatement after one day of treatment, with incremental improvements reported after each subsequent day of treatment. Some participants reported moderate improvement of symptoms during and immediately following treatment but later reported that symptoms had improved considerably beyond our scheduled follow up period.

Our most significant finding was a dramatic reduction in fNCI scores across all patients, particularly inter-region functional connectivity. All participants, regardless of self-reported symptom measures, showed a dramatic trend towards normal intra-region metabolism (t=4.72) and normal inter-region connectivity (t=7.11).

In most patients, improved fNCI scores correlated with improved self-reported symptom measures. Interestingly, some patients who showed substantial reduction in PCS symptoms immediately after treatment exhibited exacerbated symptoms 1-2 weeks post-treatment, although fNCI scores for the same patients were still significantly reduced, suggesting long-lasting metabolic and connective changes in the brain that could lead to further symptom improvements when coupled with effective therapies.

### Clinical Implications (preliminary)

Dramatic, rapid reduction in abnormal regional activation levels and inter-region connectivity demonstrates promise for fMRI-guided accelerated TMS as an intervention for chronic PCS. The presence of long-term changes, as indicated by improved fNCI scores as far as 1-month post-intervention, suggests that accelerated iTBS TMS applied to the correct location could have a normalizing effect on compensatory brain network inefficiencies typically seen in PCS patients ^9^. We propose that by correcting maladaptive neural patterns introduced by mTBI via fMRI-guided accelerated TMS, patients may then benefit from other traditional interventions for PCS to which they may have been previously unresponsive.

### Strengths

This pilot study is, to our knowledge, the first to introduce a short course accelerated iTBS protocol for treatment of chronic PCS. Our novel target identification method coupled with imaging-guided targeting for treatment led to demonstrably improved interregional functional connectivity across all patients. Objective data from task-related fMRI allows us to see functional changes in the brain attributable to our intervention.

### Limitations

Although our preliminary findings are significant, the present study has several limitations which should be considered. Firstly, we investigated a relatively small number of heterogeneous individuals, which may limit generalization of findings across broader populations. Second, this pilot study included no control groups; accordingly, all participants and administrators were aware of the treatment being delivered. Finally, our follow-up period for participants was limited and varied by a matter of weeks between some patients. Further investigation into this treatment modality should be conducted via a randomized controlled trial, with a longer follow-up period over the course of several months.

## Conclusion

Rapid accelerated fMRI-guided iTBS TMS represents a promising avenue for treatment of chronic post concussive symptoms. This intervention is non-invasive, requires relatively little time in-clinic compared to other TMS interventions, is personalized to each patient, and is well-tolerated. Our novel approach offers rapid relief from chronic concussion symptoms, and significantly reduces maladaptive inter-region connectivity issues associated with long-term PCS symptoms. Further research in a controlled clinical setting is warranted to explore the full scope, duration, and potential benefits of this novel treatment.

## Data Availability

De-identified individual participant data (including clinical scores and derived neuroimaging metrics) reported in this paper can be made available upon reasonable request directed to the corresponding author. Due to the sensitive nature of the data, requesters will need to provide a research proposal outlining the intended use of the data and obtain approval from their local Institutional Review Board (IRB) or ethics committee. A data use agreement between institutions will be required prior to data release.

## ACKNOWLEDGEMEN

We gratefully acknowledge the technical support provided by ANT Neuro throughout the use of their Visor2 neuronavigation system. Our deepest appreciation goes to the participants of this trial; their commitment and willingness to contribute to this research into post-concussive syndrome are invaluable. Special recognition is also due to Dr. Packer for her pioneering role as the first patient to successfully undergo this accelerated iTBS protocol.

## References

1. Maas AIR, Menon DK, Adelson PD, et al. Traumatic brain injury: Integrated approaches to improve prevention, clinical care, and research. The Lancet Neurology. 2017;16(12):987. doi: 10.1016/s1474-4422(17)30371-x.

2. Suwaryo PAW, Kadir F, Omar A, Singh SKD, Bolong MF. The impact of interpersonal support on quality of life in traumatic brain injury patients – a one-month post-treatment analysis. Eur J Clin Exp Med. 2024;22(3):538. doi: 10.15584/ejcem.2024.3.8.

3. Polinder S, Cnossen MC, Real RGL, et al. A multidimensional approach to post-concussion symptoms in mild traumatic brain injury. Front Neurol. 2018;9. doi: 10.3389/fneur.2018.01113.

4. Carroll LJ, Cassidy JD, Holm L, Kraus J, Coronado VG. Methodological issues and research recommendations for mild traumatic brain injury: The who collaborating centre task force on mild traumatic brain injury. Journal of Rehabilitation Medicine. 2004;36(0):113. doi: 10.1080/16501960410023877.

5. Lagarde E, Salmi L, Holm LW, et al. Association of symptoms following mild traumatic brain injury with posttraumatic stress disorder vs. postconcussion syndrome. JAMA Psychiatry. 2014;71(9):1032–1040. https://pubmed.ncbi.nlm.nih.gov/25029015/. Accessed Jul 22, 2025. doi: 10.1001/jamapsychiatry.2014.666.

6. Malenka RC, Bear MF. LTP and LTD: An embarrassment of riches. Neuron. 2004;44(1):5–21. https://www.cell.com/neuron/abstract/S0896-6273(04)00608-7. Accessed Jul 22, 2025. doi: 10.1016/j.neuron.2004.09.012.

7. Cole EJ, Stimpson KH, Bentzley BS, et al. Stanford accelerated intelligent neuromodulation therapy for treatment-resistant depression. AJP. 2020;177(8):716. doi: 10.1176/appi.ajp.2019.19070720.

8. Bouchard HC, Higgins KL, Amadon GK, et al. Concussion-related disruptions to hub connectivity in the default mode network are related to symptoms and cognition. J Neurotrauma. 2024;41(5-6):571–586. https://pubmed.ncbi.nlm.nih.gov/37974423/. Accessed Jul 22, 2025. doi: 10.1089/neu.2023.0089.

9. Anliker AE, Chauvigné LAS, Allaman L, Guggisberg AG. Neural correlates of fatigue after traumatic brain injury. Brain Commun. 2025;7(2):fcaf082. https://pubmed.ncbi.nlm.nih.gov/40066106/. Accessed Jul 22, 2025. doi: 10.1093/braincomms/fcaf082.

10. Jamieson AJ, Harrison BJ, Razi A, Davey CG. Rostral anterior cingulate network effective connectivity in depressed adolescents and associations with treatment response in a randomized controlled trial. Neuropsychopharmacology. 2022;47(6):1240–1248. https://pubmed.ncbi.nlm.nih.gov/34782701/. Accessed Jul 22, 2025. doi: 10.1038/s41386-021-01214-z.

11. Uddin LQ, Kelly AM, Biswal BB, Castellanos FX, Milham MP. Functional connectivity of default mode network components: Correlation, anticorrelation, and causality. Hum Brain Mapp. 2009;30(2):625–637. https://pubmed.ncbi.nlm.nih.gov/18219617/. Accessed Jul 22, 2025. doi: 10.1002/hbm.20531.

12. Menon V, Uddin LQ. Saliency, switching, attention and control: A network model of insula function. Brain Struct Funct. 2010;214(5-6):655–667. https://pubmed.ncbi.nlm.nih.gov/20512370/. Accessed Jul 22, 2025. doi: 10.1007/s00429-010-0262-0.

13. Seeley WW, Menon V, Schatzberg AF, et al. Dissociable intrinsic connectivity networks for salience processing and executive control. J Neurosci. 2007;27(9):2349–2356. https://pubmed.ncbi.nlm.nih.gov/17329432/. Accessed Jul 22, 2025. doi: 10.1523/JNEUROSCI.5587-06.2007.

14. Gajawelli N, Geoly AD, Batail J, et al. Increased anti-correlation between the left dorsolateral prefrontal cortex and the default mode network following stanford neuromodulation therapy (SNT): Analysis of a double-blinded, randomized, sham-controlled trial. npj Mental Health Res. 2024;3(1). doi: 10.1038/s44184-024-00073-y.

15. Epps CT, Allen MD. Discovery of therapy-targeting biomarkers for post-concussion syndrome using functional neurocognitive imaging. Brain Disord Ther. 2018;07(01). doi: 10.4172/2168-975x.1000243.

16. Lorzel HM, Allen MD. Development of the next-generation functional neuro-cognitive imaging protocol - part 1: A 3D sliding-window convolutional neural net for automated brain parcellation. NeuroImage. 2024;286. doi: 10.1016/j.neuroimage.2023.120505.

17. Klein A, Ghosh SS, Bao FS, et al. Mindboggling morphometry of human brains. PLoS Comput Biol. 2017;13(2). doi: 10.1371/journal.pcbi.1005350.

18. Hu Y, Wang J, Li C, Wang Y, Yang Z, Zuo X. Segregation between the parietal memory network and the default mode network: Effects of spatial smoothing and model order in ICA. Sci Bull. 2016;61(24):1844. doi: 10.1007/s11434-016-1202-z.

19. Yang Z, Chang C, Xu T, et al. Connectivity trajectory across lifespan differentiates the precuneus from the default network. NeuroImage. 2015;89:45. doi: 10.1016/j.neuroimage.2013.10.039.

20. Trofimova A, Smith JL, Ahluwalia V, Hurtado J, Gore RK, Allen JW. Alterations in resting-state functional brain connectivity and correlations with vestibular/ocular-motor screening measures in postconcussion vestibular dysfunction. Journal of Neuroimaging. 2021;31(2):277–286. https://onlinelibrary.wiley.com/doi/10.1111/jon.12834. doi: 10.1111/jon.12834.

